# The proportion of none infected, HSV-1 alone infected, HSV-2 alone infected and HSV-1 and HSV-2 co-infected in a cohort of Sri Lankan oral fibroepithelial polyp male patients

**DOI:** 10.1101/2024.12.17.24319140

**Authors:** Manosha Lakmali Perera, Irosha Rukmali Perera

## Abstract

**Objectives:** It is not clear whether one HSV type can protect against the other or reduce clinical manifestations of the other. This study aimed to discover the proportion of non-infected, HSV-1 alone infected, HSV-2 alone infected, and HSV-1 and HSV-2 co-infected in a cohort of Sri Lankan oral fibroepithelial polyp male patients.

**Methods:** A well-defined sub-sample included 29 FEP controls, sourced from a primary sample that accurately represents the predominant oral squamous cell carcinoma (OSCC) patients in Sri Lanka. Tissue samples were taken from frozen excisional biopsies to avoid contamination and tested for HSV-DNA using a real□time PCR assay.

**Results:** More than half 15 (51.8 %) of oral FEP patients were co-infected with HSV-1 and HSV-2. There were 03(10.3%) and 05 (17.2%) patients infected with HSV-1 and HSV-2, respectively. In contrast, 6 (20.7%) of the cohort of Sri Lankan male oral FEP patients were negative for single or co-HSV infections.

**Conclusion:** The proportion of coinfections is increasing and has now surpassed that of single HSV infections among a low-risk cohort of male patients with oral fibroepithelial polyps in Sri Lanka. Well-designed studies are needed to determine the true prevalence of the most common sexually transmitted diseases, not just in clinically suspected or symptomatic patients.

## 1. Introduction

Herpes virus infections have been present since ancient Greek times. Hippocrates (460-377 BC) described herpes simplex lesions and used the Greek word “herpes,” which means “to creep or crawl,” highlighting the spreading nature of these skin lesions. Even William Shakespeare (1564-1616) is believed to have been aware of recurrent herpes simplex lesions and their transmission. In “Romeo and Juliet,” he refers to these lesions as “blister plagues.” He writes about Queen Mab, stating, “O’er ladies’ lips, which straight on kisses dream, which oft the angry Mab with blisters plagues because their breaths with sweetmeats tainted are” [1]. Nevertheless, the evolutionary history of herpes simplex viruses (HSV-1 and HSV-2) dates back about seven million years. This conclusion was drawn from a 2017 study by researchers from Cambridge and Oxford Brookes universities, published in the journal “Virus Evolution.” The researchers analyzed DNA from the Homo lineage found in fossil records to identify potential species that could have acted as intermediaries in the transmission of HSV-2, the virus responsible for genital herpes, from our ancestors to those of chimpanzees, thus crossing the species barrier [2]. The herpes simplex viruses, types 1 and 2, have been infecting humans since before they existed as a distinct species. Unlike HSV-1, the earliest proto-humans, or hominids, did not carry HSV-2 when they split from chimpanzees about 7 million years ago. Thus, humanity avoided the risk of genital herpes for nearly 5 million years. However, between 3 and 1.4 million years ago, HSV-2 transferred from African apes to the ancestors of modern humans (Homo erectus) through an intermediate hominin species known as Paranthropus boisei, which is not directly related to humans [2].

It is worth mentioning that more than 80 distinct herpes viruses have been isolated from various animal species in the past five decades comprising the family Herpesviridae [3]. This Herpesviridae group includes herpes simplex viruses (HSV), varicella-zoster viruses, cytomegalovirus, Epstein–Barr virus, and Kaposi’s sarcoma-associated viruses, which are reported to be the aetiological agents of infectious diseases in humans [3]. HSV infections into two main types: HSV-1 and HSV-2. These infections represent a significant public health concern and economic burden, leading to considerable disability and inconvenience. The prevalence of HSV infections ranges from 50% to 60%, with many individuals remaining asymptomatic and affected worldwide [1].

Acquiring HSV infection seems a non-curable condition [4] and lifelong commitment with asymptomatic and symptomatic events life time [5] Infections with these viruses are characteristically latent and asymptomatic, but with frequent reactivations, subclinical shedding and sporadic symptomatic events [6]. Usually HSV-1 causes Gingivostomatitis herpetica or cold sores; people acquire HSV□1 through the orolabial mucosal contact with the virus in sores or saliva [7] but can also cause genital herpes [1]. Rare complications of HSV-1 infection include severe neurological, corneal, or muco-cutaneous [8].In contrast, HSV-2 is mainly transmitted by intercourse [4]. Vertical mother-to-child transmission can cause neonatal herpes, a severe and sometimes fatal disease in newborns [4]. Clinical manifestations of HSV-2 are genital ulcers [7]. HSV-2 infection may increase the acquisition and transmission of HIV [9], perhaps resulting in a synergy between these two infections. HSV infection is not a reportable disease thus; prevalence is incoherently available in global context. With the alarming finding local male non-high-risk adults (hospital in-patients) > 40 years of age had equal seroprevalence to male STD attendees of 20-29 years of age in a study conducted to find out the burden of Herpes simplex virus –type 1and 2 in Sri Lanka and 70% of HSV infections remain unrecognized [1], the present study aimed to find out asymptomatic single and co-infections due to Herpes Simplex Virus in a cohort of oral fibroepithelial polyp male patients in Sri Lanka among patients visited nine OMF units representing six, provinces in Sri Lanka.

## 2. Methods

### 2.1 Ethical approval

The study sample of this study obtained ethical approval from the Faculty Research Committee, Faculty of Dental Sciences, University of Peradeniya, Sri Lanka (FRC/ FDS/UOP/E/2014/32) and Griffith University Human Research Ethics Committee, Australia (DOH/18/14/ HREC).

### 2.2 Study design, study population and inclusion criteria

A well-defined sub-sample included 29 FEP controls, sourced from a primary sample that accurately represents the predominant oral squamous cell carcinoma (OSCC) patients in Sri Lanka. The sample size of the main study was calculated using the method proposed by Kelsey et al. [15] for unmatched case-control studies, ensuring the robustness of our findings. Consequently, this essential primary sample comprised 134 OSCC cases and 134 FEP controls, in addition to other benign mucosal lesion controls previously mentioned. For our OSCC cases, we established clear inclusion criteria: The control group consisted of Sinhalese males with clinically diagnosed fissured epithelial plaques (FEPs) involving the same anatomical sites, providing a relevant baseline for comparison and strengthening the validity of our research.

### 2.3 Study setting

Selected Oral and Maxillo-Facial (OMF) units across Sri Lanka were visited to assess the feasibility and logistics of study settings, covering six provinces: Western, Southern, Sabaragamuwa, North Western, Uva, and Central. The visits took place from November 1, 2014, to December 30, 2014. A thorough evaluation of service delivery functions was conducted on specific clinic days to understand the socio-demographic profile and clinical presentations of patients attending these clinics, particularly focusing on Oral Squamous Cell Carcinoma (OSCC) cases and the control group for Facial Erythema and Pain (FEP). This assessment included a review of biopsy reports from the past 12 months, critical observations of routine practices, and the patient turnover related to attendance patterns for OSCC and FEP cases. Based on the findings from this assessment, the inclusion and exclusion criteria were developed as outlined in section 3.8 of this chapter. The logistics and infrastructure of the OMF units, along with the dynamics of the staff, were also taken into account. Each OMF unit consisted of an OMF surgeon, house officers, nursing staff, and health assistants who actively participated in routine service provision. Biopsies were performed by house officers, while nursing staff and health assistants supported these procedures in the OMF units.

**Figure 1:**
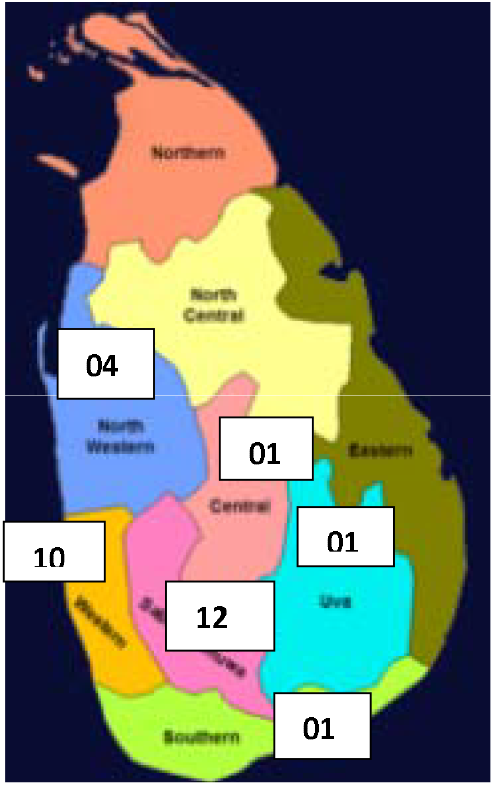
Distribution of samples by location of OMF units by Provinces in Sri Lanka

### 2.4 Study period

Study period was 17/04/15 to 02/08/15 as described previously [10].

### 2.5 Exclusion criteria

Patients who were less than 40 years of age and who had taken antibiotics in past two months previously [10].

### 2.6 Sampling technique

Non-probability consecutive sampling as described previously [10].

### 2.7 Study instruments

2.7.1 A structured pre-tested interviewer administered questioner with both open ended and closed ended questions was used to collect data on socio-demographic information, tooth cleaning practice, risk habit profile, daily consumption of fruits and vegetables, family history and awareness of oral cancer and clinical oral indicator as described previously [10].

2.7.2 Excisional biopsies of FEP were used as in this study as explained earlier [10].

### 2.8 Tissue Sampling, Genomic DNA Extraction and Quality Assessment

Deep tissue samples (∼100 mg each) were dissected from frozen (stored at □80°C) excisional biopsies to prevent contamination from the tumor surface. Genomic DNA was then extracted using the Gentra Puregene Tissue kit (Qiagen) according to the manufacturer’s protocol for solid tissue [Cat no. 158689] as described previously [10,11,12,13]. The concentration and purity of extracted genomic DNA were assessed by the nano-drop spectrophotometry. The quality assessment of the integrity and absence of PCR inhibitors was done by GAPDH (human housekeeping gene) using β□globin PCR with the primers PCO3 and PCO4 as described previously [10,11,12,13]. *Real Time PCR for HSV-1 and HSV-2 DNA*

The real time PCR assay for HSV-1 and HSV-2 were set up separately using primer sequences as standardized, validated and published previously [7,14].

#### Primers used for HSV rt PCR assay

HSV-1-wield-F CGGCGTGTGACACTATCG HSV-1-wied-R GGCGTGTGACACTATCG

Watz-HSV2-F CGCCAAATACGCCTTAGCA HSV2-R GAAGGTTCTTCCCGCGAAAT

#### Positive Controls

Extracted DNA from saliva from a patients known to be HSV-1and HSV -2 positive respectively as described previously [7].

#### Negative Controls

Extracted DNA from saliva from a patients known to be HSV-1and HSV -2 negative respectively as described previously [7].

Then, the real time PCR was performed on a Quant Studio 6-real -time machine with an initial step off-hold stage of polymerase activation step at 95^0^ C for 5 minutes, followed by 45 cycles of amplication (5seconds denaturation at 95^0^ C for 5 minutes; 30 seconds annealing (TM)/extension at 55^0^ C) and melt curve stage of 3 steps (95^0^ C for 10 minutes, 50^0^ C for 10 minutes and 95^0^ C for 15 minutes) 3. Overall run duration was 73 minutes and 24 seconds as described previously [7, 14].

#### Data analysis

Amplicon detection was determined via real time PCR screen and melt curve analysis. Positivity was determined via qPCR screening and melt curve analysis. If the melt curve temperature does not equal that of the calibration curve, that sample is reported virus-negative as described previously [7, 14]. Data entering and analysis were performed by the SPSS-21, Statistical Package. The statistical significance of pre tested interviewer administered data was obtained by descriptive statistics for percentage, frequency, mean, and standard deviation.

## 3. Results

The results the proportion of none infected, HSV-1 alone infected, HSV-2 alone infected and HSV-1 and HSV-2 co-infected in a cohort of Sri Lankan oral fibroepithelial polyp male patients, a subset of a main unmatched case-control study comprised of 134 cases with oral cancer and 134 controls with benign mucosal lesions presented here. This subset consisted of 29 Sinhalese males having OSCC in the buccal mucosa and tongue. The socio-demographic profile of cases and controls is described below.

### 3.1 The socio-demographic profile of FEP patients

Table 01 provides an overview of the socio-demographic profile of the first-episode psychosis (FEP) subjects. The mean age of the participants was 49.96 years, with a standard deviation of 13.38 years. Of them, the largest group, comprising 11/29 (37.9%) of the FEP patients, had completed education beyond secondary school, followed by 10/29 (34.5%) who had attained secondary education and 7/29 (24.1%) who had only completed primary education. The minority, accounting for1/29 (3.5%), reported having no formal schooling. Overall, a higher proportion of the study subjects had educational attainment at the secondary level or above. So, the clerical/professional category was 12/29 (41.4%), and 8/29 (27.6%) was skilled/unskilled manual categories. The proportion of farmers was 9/29 (31.0%).

**Table 01.**
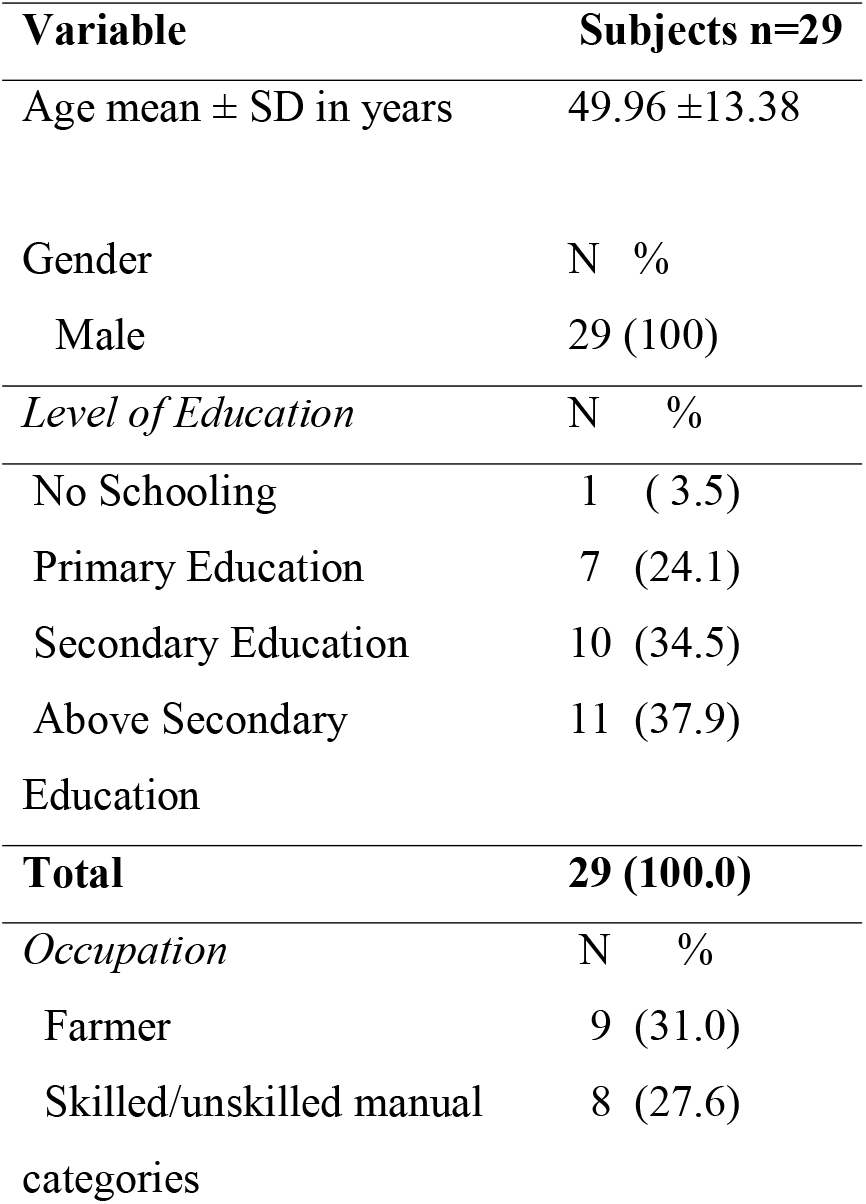
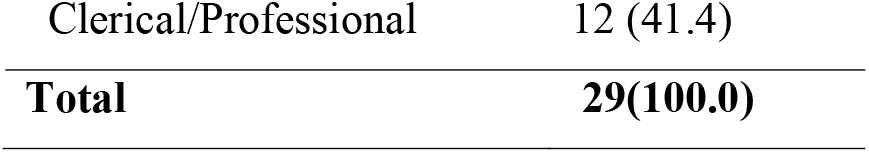
Distribution of study subjects by level of education and occupation

| **Variable** | **Subjects n=29** |  |
| --- | --- | --- |
| Age mean ± SD in years | 49.96 ± 13.38 | |
| Gender | N | % |
| Male | 29 | (100) |
| *Level of Education* | N | % |
| No Schooling | 1 | ( 3.5) |
| Primary Education | 7 | (24.1) |
| Secondary Education | 10 | (34.5) |
| Above Secondary Education | 11 | (37.9) |
| **Total** | **29 (100.0)** |  |
| *Occupation* | N | % |
| Farmer | 9 | (31.0) |
| Skilled/unskilled manual categories | 8 | (27.6) |

|  |  |
| --- | --- |
| Clerical/Professional | 12 (41.4) |
| **Total** | **29(100.0)** |

**Table 1:** Distribution of oral FEP patients by categories of HSV infections

| Categories of HSV infections | Oral FEP patients |  |
| --- | --- | --- |
|  | No | % |
| None infected | 06 | 20.7 |
| HSV-1 alone infected | 03 | 10.3 |
| HSV-2 alone infected | 05 | 17.2 |
| HSV-1 and HSV-2 co-infected | 15 | 51.8 |
| Total | 29 | 100.0 |

### 3.2 Distribution of oral FEP patients by none infected, HSV-1 alone infected, HSV-2 alone infected and HSV-1 and HSV-2 co-infected in a cohort of Sri Lankan male patients

## 4. Discussion

To the best of the authors’ knowledge, this is the first time that the presence of the highest proportion of 15/29 (51.8%) of HSV-1/2 coinfections and the detection in asymptomatic, thus low-risk patients attended to OMF units with clinically diagnosed FEP have been revealed in Sri Lanka. Thus, there is a lack of information on similar studies to compare and contrast with. The finding of HSV 1/2 co-infections was more than twofold higher than the previous finding of 20% of >50 years old clinically suspected in a study by Fernando and colleagues [15]. When it comes to regional context, the overall seroprevalence of HSV□1 singly infected, HSV□2 singly infected, and both infected but not HIV infected was 71.18%, 5.08%, and 3.38% without genital ulcers disease (GUD) respectively, a study from a tertiary care setting India is in contrast to the findings of the present study [16]. In a study conducted to find out socio-demographic and behavioral correlates of herpes simplex virus type 1 and 2 infections and co-infections among adults in the USA, it has been detected Approximately 48% were positive for HSV-1 alone, 7% were positive for HSV-2 alone and 12% were co-infected with HSV-1 and HSV-2 [4]. Thus, the HSV 1/2, co infections were four fold higher in our study and HSV -2 alone was higher than HSV-1 alone in the present study [4].

In our study cohort, the proportion of asymptomatic HSV-1 infections among males with a mean age of 49.96 ±13.38 was 03/29 (10.3%). Our finding relies on the seroprevalence (HSV□1 IgG positive) of 4% and 12 % of Sri Lankan older adults 50 years and>50 years reported previously [15]. Moreover, 05/29 (17.2%) of asymptomatic patients were infected by HSV-2 alone in our study. This means the proportion of asymptomatic HSV-2 was higher than HSV-1. This finding is contradictory with the findings of 79% of HSV-1 but 11% of HSV-2 sero positivities among Sri Lankan volunteer blood donors attended to North Colombo Teaching Hospital Ragama [16], HSV-1 (37% and 60%) was higher than that of HSV-2 (3% and 20%) seroprevalence in 15-50years and >50years old clinically suspected individuals respectively [15] as well as of 58.8 % (49.4–67.8, 95% CI) CI of HSV-1 antibodies compared with HSV-2 antibodies 10.1% (5.3–17, 95% CI) amongst Sri Lankan migrant workers in Qatar [17]. However, the present study corroborates the finding of the adjusted odds ratios of HSV-2 antibody sero positivity of Sri Lankan males, who were non attendees of sexually transmitted diseases (STD) clinics was (95% CI) 1.21 (0.96 to 1.60) and significantly higher HSV-1 prevalence than Sri Lanka (p<0.000) after adjusting for age in another study [18].

In the global context, an estimated 3.7 billion people under age 50 (67%) have HSV-1 infection, concerning genital HSV-1 infection, 140 million people aged 15-49 years were estimated to have genital HSV-1 infection worldwide in 2012 [19]. Genital herpes caused by HSV-2 is a global issue, and an estimated 417 million people worldwide were living with this in 2012 [19]. In the Middle East, one sero-epidemiology study based on asymptomatic university students attending Islamic Azad University of Kazeroun, southwest of Iran disclosed that HSV-1 and HSV-2 IgG antibodies were positive in 285 (79.2%) and 84 (23.3%) subjects, respectively. The seroprevalence of HSV-2 was higher among females (29.0%) compared with males (17.5%) (p<0.05), however, there was no significant correlation between gender and HSV-1 seroprevalence [20]. In the USA, it is as high as 40-63% is seropositive for HSV-1 and 16-18% is seropositive for HSV-2 than other European countries but lower than in Sub-Saharan Africa [21]. This finding was not consistent with our finding of more HSV-2 alone than HSV-1 alone [4].

Our findings challenge the widely held belief that the most common method of horizontal transmission of HSV-1 in childhood is stigma-free as HSV-1 acquisition probably reflects extensive infant salivary spread in poorer hygienic conditions [20]. However, our study supports the recognition of HSV-1 as a cause of first-episode genital herpes in various countries [21]. Currently, there is insufficient data regarding the shedding of HSV into the oral cavity during the prodromal stage of the disease, recurrences, and asymptomatic periods, as indicated by the excisional biopsies of first-episode patients. Sero-epidemiological studies are needed to better understand the pattern and distribution of HSV infection within populations, given that up to 70% of genital herpes infections go unrecognized and most infected individuals intermittently shed the virus, making them infectious [18]. According to unpublished data of Prof. Anura Weerasinghe herpes simplex is like cough and cold in Sri Lanka. Thus, our study provides alarming findings on this sexually transmitted disease. On one hand, HSV-1 epidemiology is transitioning in Asia. HSV-1 is probably playing a significant role as a sexually transmitted infection, explaining one-fifth of genital herpes cases. On the other hand, sexual activity not only played an import role for HSV-2 and it is equally important for HSV-2 when considering the highest proportion of 15/29 (51.8%) of HSV-1/2 coinfections and the detection in asymptomatic, thus low-risk patients attended to OMF units with clinically diagnosed FEP have been revealed in Sri Lanka. Before public health interventions it is of utmost importance to know the disease burden of HSV not only assessing the quarterly return of sexually transmitted disease including HIV/AIDS for epidemiological bulletin of the epidemiology unit of Ministry of Health Sri Lanka. Small sample size was a limitation of our study.

## 5. Conclusion and recommendations

The proportion of coinfections is rising and surpassing single HSV infections in a low-risk cohort of Sri Lankan oral fibroepithelial polyp male patients. Additionally, HSV-2 infection is more prevalent than HSV-1 in the same asymptomatic cohort. Transmission of an HSV infection is possible symptomatic or asymptomatic reactivation, which can be either spontaneous or triggered by external stimuli and high-risk behaviors of transmission and is geographic and population-specific. Thus, descriptive cross-sectional studies with large sample sizes and controlling for confounders to assess the prevalence of HSV infections and associated risk factors among high-risk and low-risk populations and awareness programs on safe sex practices and avoidance of infectious routes are much recommended.

## Data Availability

All data produced in the present study are available upon reasonable request to the authors

## AUTHOR CONTRIBUTIONS

### Manosha Perera

Conceptualization; experimenta design; laboratory analysis; interpretation of results obtained by laboratory and statistical analysis; writing the original draft.

### Irosha Perera

Conceptualization; study design; sample size calculation; performing excisional biopsies; followed patients and revision; statistical analysis; revision of the manuscript.

## CONFLICT OF INTEREST STATEM ENT

The authors declare no conflict of interest.

## ETHICS STATEMENT

The microbiome profile of oral squamous cell carcinoma tissues in a group of Sri Lankan male patients which received ethical approval from the Faculty Research Committee, Faculty of Dental Sciences, University of Peradeniya, Sri Lanka (FRC/ FDS/UOP/E/2014/32) and Griffith University Human Research Ethics Committee, Australia (DOH/18/14/ HREC).

## ACKNOWLEDGEMENT

We acknowledge the late Professor Newell Johnson and Associate Professor Glen Ulett, Professor of Microbiology at the School of Medical Science and Pharmacy, Gold Coast Campus, Griffith University, for their valuable contributions that helped make this study a success. We are grateful to Professor W.M. Tilakaratne, Senior Professor of Oral Pathology, and Professor L. Samaranayake, Professor of Oral Microbiology, for their guidance throughout this research. We also thank the Oral and Maxillofacial Surgeons — Dr. Sharika Gunathilake, Dr. S.A.K.J. Kumara, Dr. Ranjith Lal Kandewatte, Dr. P. Kirupakaran, Dr. D.K. Dias, Dr. Chamara Athukorale, Dr. Suresh Shanmuganathan, and Dr. T. Sabesan — for facilitating data and sample collection from their respective Oral and Maxillofacial Units. Last but certainly not least, we express our appreciation to Dr. Rohitha Muthugala, Consultant Virologist at the National Hospital, Kandy, Sri Lanka, for his crucial advocacy role.

